# Clinical Features Associated with COVID-19 Outcome in MM: First Results from International Myeloma Society COVID-19 Dataset

**DOI:** 10.1101/2020.08.24.20177576

**Authors:** Ajai Chari, Mehmet Kemal Samur, Joaquin Martinez-Lopez, Gordon Cook, Noa Biran, Kwee Yong, Vania Hungria, Monika Engelhardt, Francesca Gay, Ana García Feria, Stefania Oliva, Rimke Oostvogels, Alessandro Gozzetti, Cara Rosenbaum, Shaji Kumar, Edward A. Stadtmauer, Hermann Einsele, Meral Beksac, Katja Weisel, Kenneth C. Anderson, María-Victoria Mateos, Philippe Moreau, Jesus San-Miguel, Nikhil C. Munshi, Hervé Avet-Loiseau

## Abstract

The primary cause of morbidity and mortality in patients with multiple myeloma (MM) is an infection. Therefore there is great concern about the susceptibility to the outcome of COVID-19 infected patients with multiple myeloma.

This retrospective study describes the baseline characteristics and outcome data of COVID-19 infection in 650 patients with plasma cell disorders (98 outpatinets and 538 hospitilized patinets), collected from 10 countries by the International Myeloma Society to understand the initial challenges faced by Myeloma patients during COVID-19 pandemic. Descriptive statistics, univariate logistic regression, and multivariate analysis were performed for hospitalized MM patinets.

The median age was 69 years, and nearly all patients (96%) had MM. Approximately 36% were recently diagnosed (2019-2020), and 54% of patients were receiving first-line therapy. Thirty-three percent of patients have died, with significant geographic variability, ranging from 27% to 57% of hospitalized patients. Univariate analysis identified age, ISS3, high-risk disease, renal disease, suboptimal myeloma control (active or progressive disease), and one or more comorbidities as risk factors for higher rates of death. Neither history of transplant, including within a year of COVID-19 diagnosis nor other anti-MM treatments were associated with outcomes. Multivariate analysis found that only age, high-risk MM, renal disease, and suboptimal MM control remained independent predictors of adverse outcome with COVID-19 infection.

The management of MM in the era of COVID-19 requires careful consideration of patient and disease-related factors to decrease the risk of acquiring COVID-19 infection, while not compromising the disease control through appropriate MM treatment. This study provides the data to develop recommendations for the management of MM patients at risk of COVID-19 infection.

**Key Points:**

- High but variable mortality for hospitalized MM patients (27% to 57%)
- Optimal MM control was associated with COVID-19 associated death for MM patinets

**Explanation of novelty:** This study investigated the risk and outcome of COVID-19 infection in MM patients globally (10 countries)

## INTRODUCTION

In the current severe acute respiratory syndrome coronavirus 2 (SARS-CoV-2)^1^ pandemic, known as COVID-19, cancer represents a major risk factor^2-4^ for COVID-19 associaed death. Cancer patients with COVID-19 represented 8.3% of deaths in the New-York city area, and 7.6% deaths from the Wuhan area in China.^5,6^ An even higher fatality rate (20.3%) was observed in Italy^7^. A recent study aimed at identifying risk factors for death in cancer patients developing COVID-19 interestingly (but not surprisingly) reported age over 65 as a risk factor. In this study, treatment with checkpoint inhibitors was a risk factor, but not ongoing chemotherapy^5^.

Multiple myeloma (MM) is a hematological cancer involving plasma cells, mostly within the bone marrow^8^. Apart from the specific cancer-related symptoms, most patients display immunosuppression^9^, involving both the B cell and T cell compartments. Infections are a common disease complications^10^, and unfortunately remain a major cause of death. Furthermore, corticosteroids, and especially dexamethasone, are used as treatment throughout the disease course, usually at high doses^11,12^. This MM therapy may increase the immunosuppression observed in patients with MM, though low doses seem to improve mortality in hospitalized patients. MM usually affects the elderly population, a more vulnerable group of patients due to immunosenescence together with other comorbidities. In addition, the younger MM patients are usually treated with high dose chemotherapy followed by autologous stem cell transplant (ASCT)^13^, with high infection susceptibility during the 3-month period following transplant^14^. For all these reasons, MM could theoretically represent a high-risk factor for poor outcomes with COVID-19^15^.

In this international study, we have collected data and investigated the risk and outcome of COVID-19 infection in MM patients globally, both to evaluate the death rate and to identify potential risk factors that could be modified to improve patient outcomes during the current pandemic and in future. With this in mind, we have predominantly focused our analysis on patients requiring hospitalization for COVID-19infection.

## METHODS

### Patient Cohort

The International Multiple Myeloma COVID-19 dataset created by the International Myeloma Society (IMS) has retrospectively collected data for 650 patients with a plasma cell disorder from ten different countries and multiple centers. All patients in the study had confirmed positive SARS-CoV-2 tests, according to the protocols in their respective countries. A questionnaire created by IMS was shared with participant institutes/investigators, and all required information was collected by the participating investigators. Data cleaning, pre-processing, and quality control was completed before the data analysis. COVID-19 outcome is defined as recovery from the virus and discharge from hospital or death due to COVID-19. Those patients who required an ongoing treatment at the time of data collection, unknown diagnosis or unknonw COVID-19 outcomes and not hospitilized were excluded from the cohort for statistical analysis.

### Statistical Analysis

All statistical analyses were performed using R. Descriptive statistics for demographic information and clinical variables are reported. Parametric two group comparison was used for age, univariate logistic regression was used to evaluate the association between COVID-19 outcome and variables, and odds ratios (OR) with 95% confidence intervals (CI) were estimated. Multivariate analysis was performed using only variables that were associated with the outcome on univariate analysis.

## RESULTS

Overall 650 patients with a plasma cell disorder and COVID-19 infection are included in this study, with the majority of patients from Spain (28.62%), France (28.46%), USA (19.38%), and the United Kingdom (14.77%) (Table 1, Figure 1A, Supplementary Table 1). Median age was 69 years (Range 34-92 years), and 58.5% of patients were male (Table 2). The vast majority of patients (95.5%) had MM, while 29 (4.5%) patients had another plasma cell disorder. The MM immunoglobulin subtype included 55% IgG, 21% IgA and 20% light chain(Table 2). Patients were equally distributed between ISS stages 1-3, with 32.1% patients having high-risk cytogenetics and 26.5% with renal dysfunction. Fifty-four percent of patients were receiving first line of therapy, while 23.5% of patients had three or more lines of therapy. Additional demographic data are presented in Table 1, Table 2, and Supplementary Tables 1 and 2.

**Table 1:**
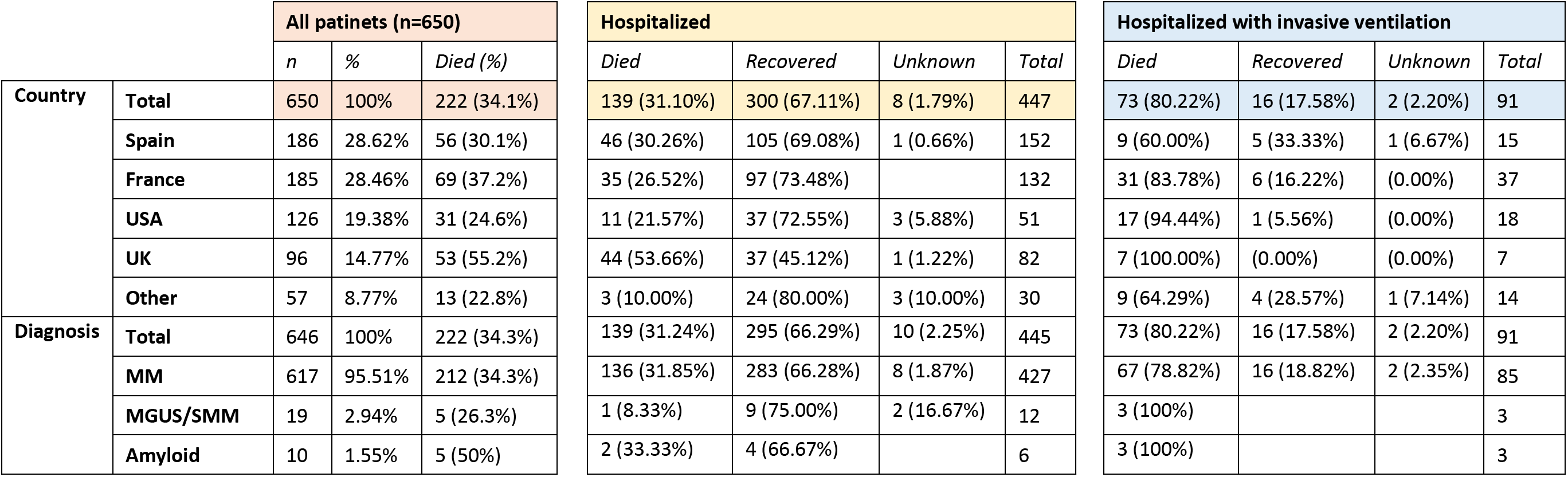
Total number of patients and their COVID-19 outcomes recorded in the International Myeloma Society COVID-19 dataset by country and diagnosis. (All patinets (n=650) refers to all the patinets including, hospitalized and outpatients without any exclusion, in our dataset.

**Figure 1:**
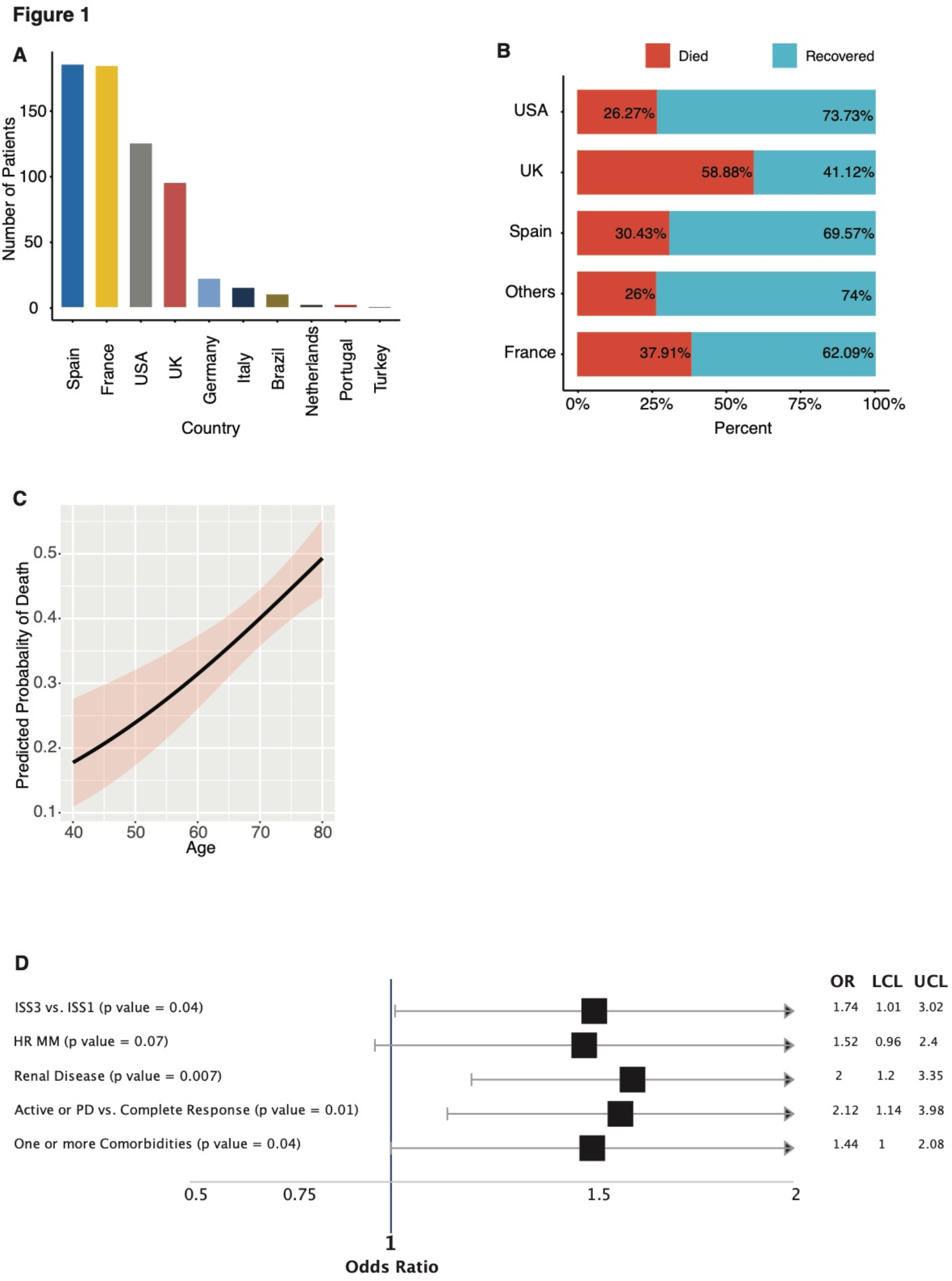
**A)** Number of patients in the IMS COVID-19 dataset with plasma cell disorders. **B)** Overall (outpatient and hospitalized) COVID-19 death rates in the dataset by contributing countries. **C)** Predicted COVID-19 outcome for MM patients by age. **D)** A forest plot for risk factors for MM patients from univariate analysis (OR, odds ratio; UCL, upper confidence level and LCL, lower confidence level).

**Table 2:**
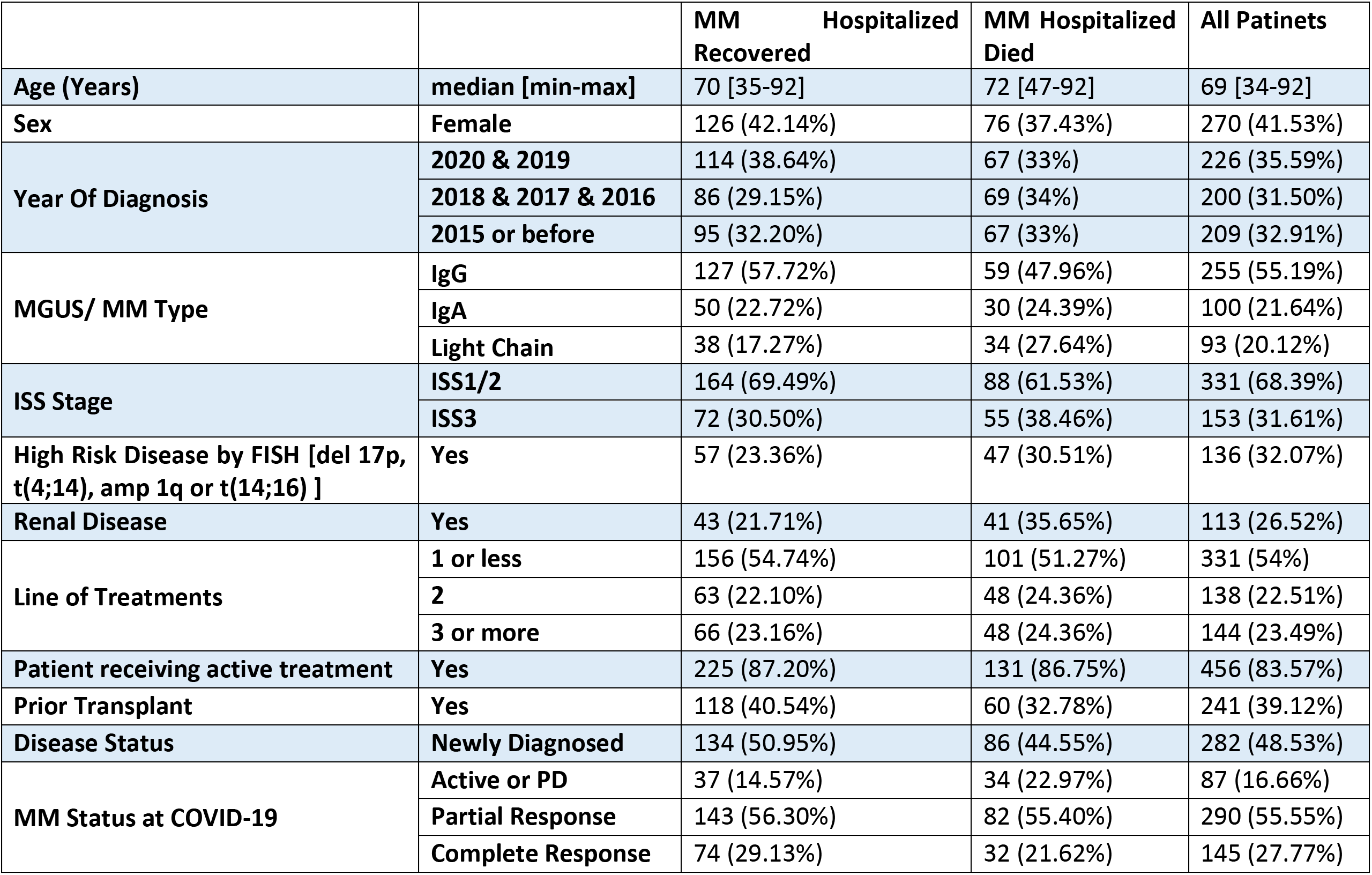
Patient characteristics for hospitalized MM patients and overall dataset.

Thirty three percent of patients died following COVID-19 diagnosis. Death rate increased from 4% for those who were outpatients to 31% for hospitalized patients not on ventilator support to 80% for patients on ventilator support (Table 1 and Supplementary Table 1). The variability in death rates across 4 major countries is shown in Figure 1B and Table 1. The death rate in patients with other plasma cell disorders was 31% (9 of 29).

We have further focused on analyzing the hospitalized patients, where the mortality rates ranged from 27% in Germany, Italy, Brazil, Netherlands, Portugal and Turkey, to 57% in the United Kingdom (Table 2). Age was significantly associated with COVID-19 outcome (p value < 0.001). The estimated probability of death for 40, 60, and 80 year old patients was 17.76%, 31.43%, and 49.3%, respectively (Figure 1C). Forty percent of hospitalized patients were female, and in contrast to prior reports, sex was not associated with outcome. Of note, mean age for male patients (69 years) was significantly lower than female patients (71.5 years) (p value = 0.01).

Of the patients with available data, those diagnosed with MM in 2019 or 2020 accounted for 35.6% of the cohort, and those with ≤1 line of therapy accounted for 54% of the cohort, while 32.9% of patients were diagnosed on or before 2015. Neither time from diagnosis nor number of prior lines of treatment had any impact on outcome of COVID-19 infection. Immunoglobulin (Ig) type distribution was similar to the general MM population, and isotypes were not associated with outcome. Univariate analysis identified ISS3 vs. ISS1 (p value = 0.04), high risk disease [del 17p, t(4;14), amp 1q or t(14;16)] (p value = 0.07), renal disease (p value = 0.007), inadequate MM control [active disease or progressive disease vs. complete response] (p value = 0.01), and one or more comorbidities (p value = 0.04) correlating with higher rates of death (Figure 1D).

Eighty-seven percent patients who had a known treatment status were on active MM therapy at the time of COVID-19 diagnosis, and 89% patients had their therapy held during COVID-19 diagnosis and management (Table 2 and Supplementary Table 2). A history of prior transplant or transplant within a year of COVID-19 diagnosis did not impact outcome. In fact, patients with a history of SCT within a year of COVID-19 diagnosis had a lower death rate; however, this difference was cofounded by a ten year age difference between transplant and non-transplant patients, and was not observed when adjusted for age. Similarly, we did not observe any significant difference in outcome from COVID-19 infection whether patients underwent transplant within 6 months or more than 6 months before their COVID-19 diagnosis. Approximately 86% of patients had prior exposure to proteasome inhibitors (PIs), 80% to immunomodulatory (IMiD) agents, and 30% to anti-CD38 antibody. In univariate analysis, prior PI, IMiD or anti-CD38 treatment were not associated with outcome. Although, univariate analysis showed that patients who were receiving IMiD treatment at the time of Covid-19 diagnosis had decreased mortality compared to patients not on IMIds, multivariate analysis failed to identify IMiD or any of these features as being related with outcome (Table 3). We did not observe any significant correlation between active PI, IMiD, anti-CD38 mAb, alkylating agent, steroids, or other (venetoclax, 96 hours infusional regimens, bispecific T cell engagers, belantamab, CART, elotuzumab, HDAC) treatments and the COVID-19 outcome.

**Table 3:**
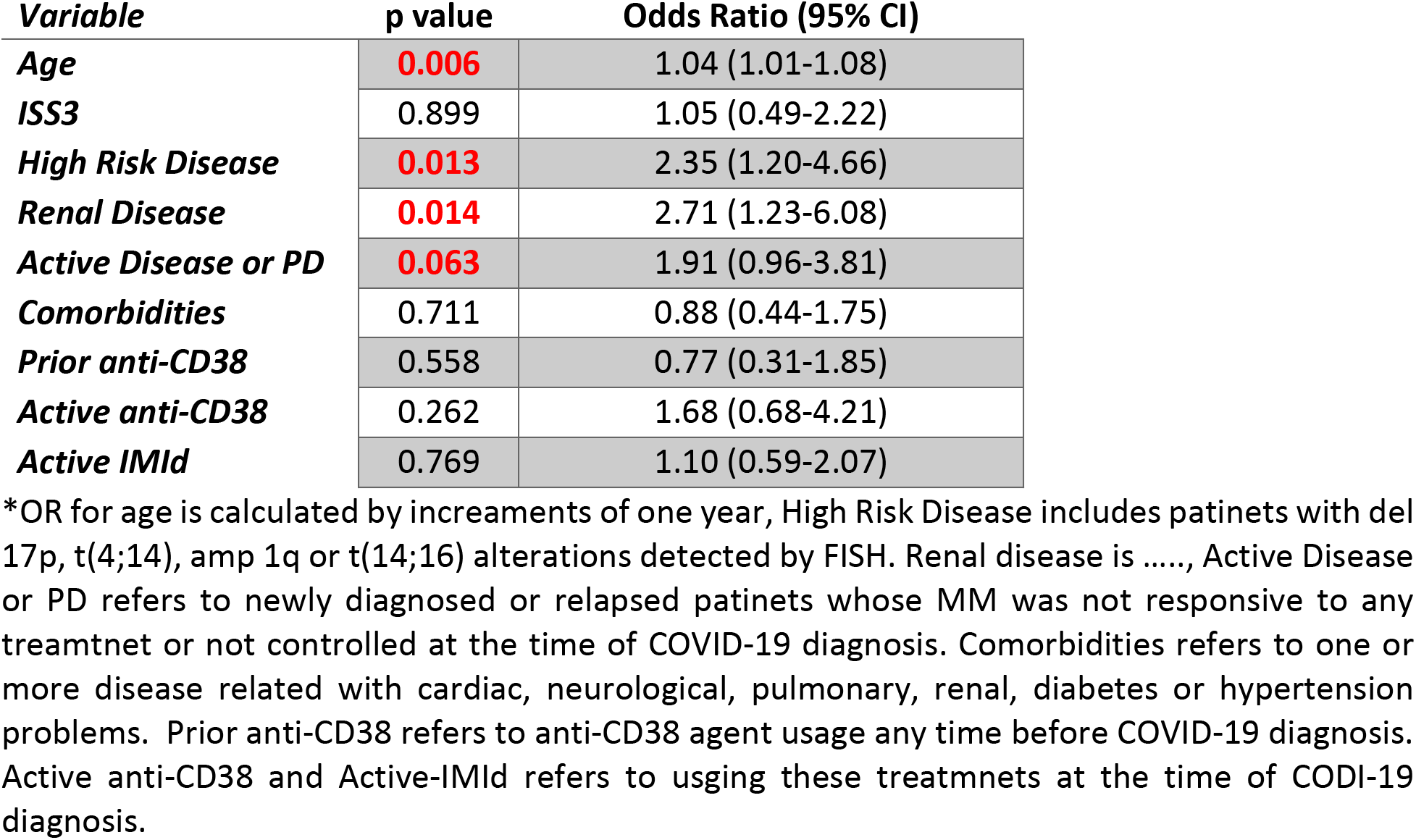
Estimated COVID-19 outcome predictors based on multivariate analysis and their odds ratios for Multiple Myeloma patients.

| **Variable** | **p value** | **Odds Ratio (95% CI)** |
| --- | --- | --- |
| **Age** | **0.006** | 1.04 (1.01-1.08) |
| **ISS3** | 0.899 | 1.05 (0.49-2.22) |
| **High Risk Disease** | **0.013** | 2.35 (1.20-4.66) |
| **Renal Disease** | **0.014** | 2.71 (1.23-6.08) |
| **Active Disease or PD** | **0.063** | 1.91 (0.96-3.81) |
| **Comorbidities** | 0.711 | 0.88 (0.44-1.75) |
| **Prior anti-CD38** | 0.558 | 0.77 (0.31-1.85) |
| **Active anti-CD38** | 0.262 | 1.68 (0.68-4.21) |
| **Active IMiD** | 0.769 | 1.10 (0.59-2.07) |

\*OR for age is calculated by increments of one year, High Risk Disease includes patients with del 17p, t(4;14), amp 1q or t(14;16) alterations detected by FISH. Renal disease is ....., Active Disease or PD refers to newly diagnosed or relapsed patients whose MM was not responsive to any treatment or not controlled at the time of COVID-19 diagnosis. Comorbidities refers to one or more disease related with cardiac, neurological, pulmonary, renal, diabetes or hypertension problems. Prior anti-CD38 refers to anti-CD38 agent usage any time before COVID-19 diagnosis. Active anti-CD38 and Active-IMiD refers to using these treatments at the time of COVID-19 diagnosis.

The treatments of COVID-19 were very heterogeneous, with the most frequent therapies including combination strategies (70%), antibiotics (14%), and hydroxychloroquine (10%). No therapies for COVID-19 appeared to be protective or associated with worse outcomes.

Of the aforementioned variables that were associated with an increased risk of death by univariate analysis, only age (OR = 1.04, 95% CI 1.01-1.08), high risk MM [del 17p, t(4;14), amp 1q or t(14;16)] (OR = 2.35,95% CI 1.20-4.66), renal disease (OR = 2.71, 95%CI 1.23-6.08) and active or progressive MM (OR = 1.91, 95% CI 0.96-3.81) remained as independent predictors of adverse outcome on multivariate analysis (Table 3).

## DISCUSSION

The COVID-19 infection has affected patients globally, with high incidence in Europe and the Americas. The disease has involved patients at all age-groups, however, heterogeneity in outcome of COVID-19 infection has been observed associated with co-morbidities, racial differences, as well as individual characteristics such as smoking^16-18^. Of note, the presence of co-morbidities has been extensively studied to identify patients at greater risk of infection and those with worse outcome. In this regard, our current study focuses on a single type of cancer, MM, to understand both impact and outcome of patients when they develop COVID-19 infection. As MM patients have hallmark immunosuppression, it is of great interest to understand impact of both disease and its treatment, ie the immunosuppressive effects of high-dose therapy with autologous transplantation, as well as novel targeted therapies.

Here we report data primarily from four countries (Spain, France, UK and USA) having high prevalence of COVID-19 infection and with the highest frequencies of COVID-19 in MM patients Differing access to testing likely led to the majority (55%) of outpatients coming from USA.

Interestingly, recent data from NYC institutions showed that approximately 19% of 127 patients with COVID-19 actually had MM precursor conditions (plasmacytoma, monoclonal gammopathy of undetermined significance (MGUS), smoldering MM, SMM)^19^. Based on the fact that data collection was feasible predominantly in patients who were hospitalized, our study has focused on hospitalized patients and their outcomes and are unable to provide insight into a question regarding susceptibility and outcome of COVID-19 in patients with MM precursor conditions.

According to SEER data, 22% of the patients with myeloma had their diagnosis of MM in 2019-20. In our cohort, 36% of the patients with COVID-19 infection were diagnosed with MM in 2019-20, suggesting higher susceptibility in earlier stages of the disease. Even if we account for variability in diagnosis and selection of cases, there seems to be no evidence of increased hospitalizations primarily in advanced multiply relapsed MM, as it was initially postulated. However, different rate of access to the testing or hospital care between newly diagnosed and established patinets may have impact on these results. Of note, a prior report noted that hypogammaglobulinemia (IgG < 700 mg/dl) was not associated with outcomes, but severe hypogammaglobulinemia (IgG <400 mg/dl) was associated with death^20^. The impact of disease stage and clinical immunoparesis is being evaluated in prospective studies. The sex distribution in this analysis was similar to general incidence of MM, and thus suggests a similar susceptibility to COVID-19 infection between male and female patients with MM. A clear association between age and outcome was confirmed in these patients, as is true in other settings.

The differences reported here between various countries reflects, at least in part, differing diagnostic and management practices, as well as the resources available and utilized in management of COVID-19 at the height of the pandemic, as well as referral patterns. Health system differences between countries may influence ability to seek or obtain SARS-CoV-2 testing and heatlh care, including admission to hospital. Healthcare provides should always consider local situations and COVID-19 positivity rate when results and recommendations made here are used as reference. For example, the number of patients who received ventilator support differed from 7% to 31%, and there are also differences in outpatient management versus hospitalization. Nonetheless, patients hospitalized can be considered to have more severe COVID-19 related complications requiring more intensive support.

Our data suggests higher mortality in hospitalized patients with MM and Covid19 infection than non-myeloma patinets. A recent study from Spain found higher mortality rate in MM patients with COVID-19 (34%) compared to age and sex matched non-MM patients with COVID-19 (23%)^21^. A recent publication from France confirmed overall mortality in all hospitalized patients with COVID-19 to be 16% which is significantly lower then mortality observed here in patients from France (39%)^22^. Our data clearly suggests a lack of relationship between prior lines of therapy, prior type of therapy, or receiving active MM therapy at the time of diagnosis with COVID-19 and outcome. Interestingly, neither past nor recent high-dose therapy had significant impact on outcome. These data indicate that it may not be necessary to postpone indicated MM therapies, including high-dose therapy, during the COVID-19 pandemic. Within the limitation of our sample size and retrospective nature of the study, there is no clear suggestion for need to avoid any specific MM treatment. Importantly, patients with good MM control (complete response, CR) had superior outcome compared to those with relapsed disease or partial response (PR). A similar finding was observed in a study of 928 cancer patients with COVID-19 where patients with active cancer (progressing vs remission) had an adjusted odds ratio of 5.2 for 30 day mortality but there was no association with recent non-cytotoxic therapy nor recent cytotoxic systemic chemotherapy^2^. As most patients receive dexamethasone as part of combination therapy, it was not possible to judge its independent impact on outcome. This is important since a recent report suggests superior survival for those COVID-19 patients given dexamethasone as part of their COVID-19 therapy^23^.

Our multivariate model identifies age, high risk or progressive MM, and presence of renal disease as indicators of poor outcome. MM therapy to achieve deep response may therefore also protect patients from adverse outcome from COVID-19 infection. While little is known about the recovery of patients with MM from COVID-19 infection, Wang et al found that the median time to PCR negativity was 43 (range 19-68) days from initial positive PCR^24^. Interestingly, 96% (22/23) of MM patients developed antibodies to SARS-CoV-2 at a median of 32 days after initial diagnosis.

Based on the observations reported here, young patients with high risk and/or active MM need to receive therapies to control their disease, which will also improve their outcome, if infected with COVID-19. For elderly patients with higher death rate from COVID-19, disease control is also beneficial, but may be achieved using regimens that decreases frequency of office visits (e.g. oral drug) in order to avoid exposure to the virus. Importantly, continued therapy including steroids and high-dose therapy are not contraindicated, and in fact should be continued to achieve better MM control, which is associated with improved outcome even with COVID-19 infection.

In conclusion, this large collaborative international effort provides the first large scale analysis and IMS initial suggestions on the management and outcomes for patients with MM during the current COVID-19 pandemic (Table 4). As the pandemic and data accumulation moves rapidly we need prospective studies on treatment options and additional patient characteristics to further understand the variables associated with COVID 19 associated death for MM patinets. Given the high mortality noted in MM patients, it highlights the critical importance of measures to prevent contracting Covid-19, such as social distancing and wearing masks, in patients with MM.

**Table 4:**
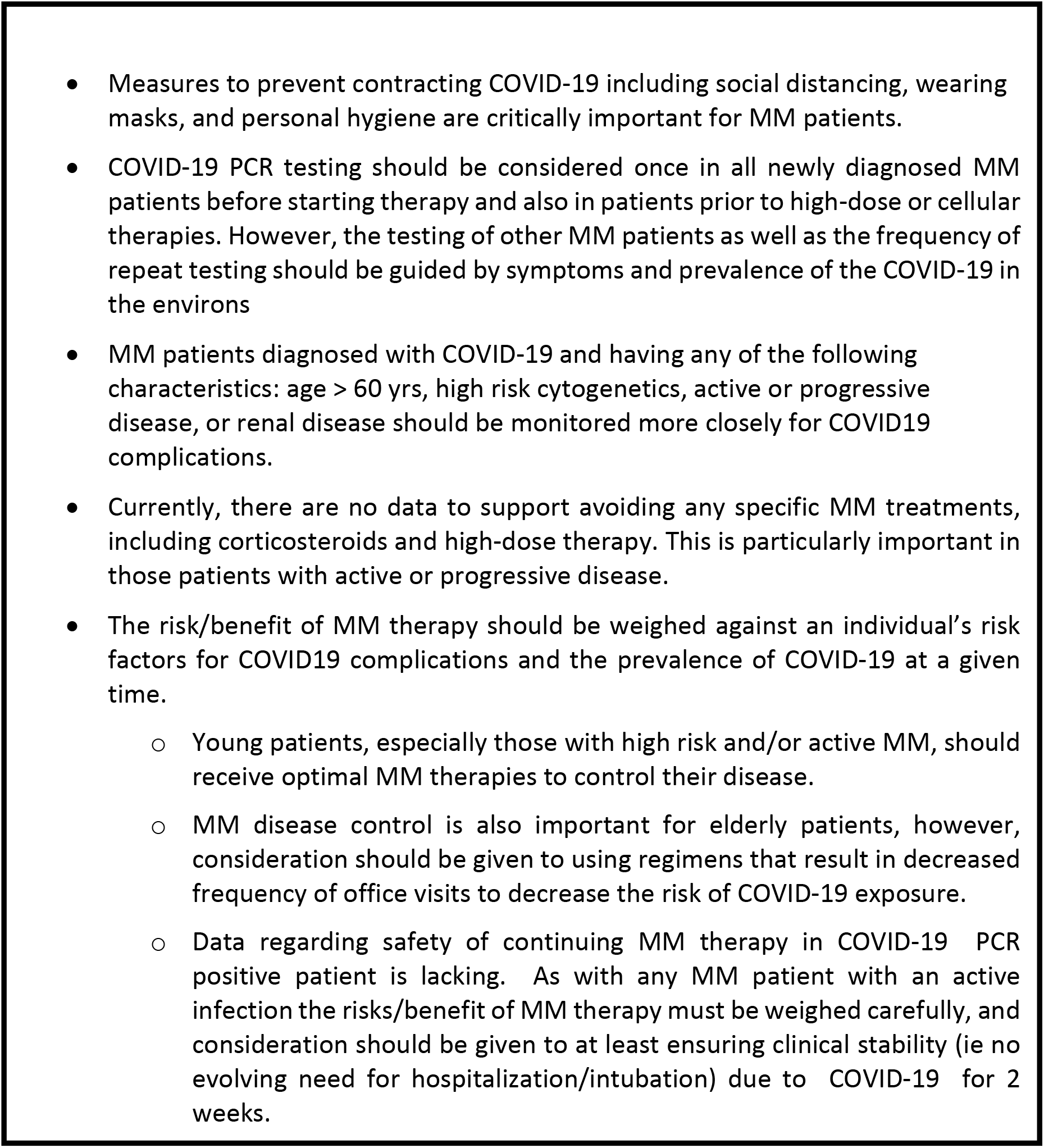
Recommendations for management of multiple myeloma patients in the era of a global COVID-19 pandemic

## Data Availability

All data used in the study can be obtained from International Myeloma Society.

## Author Contribution

AC, MKS, JSM, NCM, HAL designed the research analyzed the data. All authors contributed to data collection and wrote the manuscript.

## Acknowledgements

This work was supported by the efforts of the International Myeloma Society and its members globally. We would like to thank all medical centers and health workers for their fight against the global pandemic.

